# Clinical Information Needs Among Latin American Physicians: A Multi-Country Analysis of Semantic Clinical Search

**DOI:** 10.64898/2026.06.26.26356340

**Authors:** Katherine Monsalve, María Camila Villa, Natalia Castaño-Villegas, José Zea, Laura Velásquez

## Abstract

**Background:** Physician clinical information-seeking behavior is poorly characterized in Latin America.

**Objective:** To describe query topic patterns, geographic distribution, and trends among Latin American physicians using a semantic clinical search platform.

**Methods:** Retrospective query-log analysis, May 2025–June 2026; 9,443 physicians across 20 countries, 235,803 queries; a 10-category taxonomy applied via prefix matching.

**Results:** Activation rate: 84.4%. Management Plan (35.6%), Differential Diagnosis (20.6%), and Work-up and Test Selection (11.4%) accounted for 67.6% of queries; this hierarchy was consistent across 17 countries. Per-physician volume was broadly similar (mean 29.6 queries).

**Conclusion:** Query behavior in this Latin American physician cohort was oriented toward clinical decision support, with a consistent hierarchy across countries. Future studies should evaluate clinical impact.

## Introduction

The volume of biomedical knowledge has grown beyond what any individual clinician can routinely monitor, creating sustained demand for clinical information retrieval tools. Clinical decision support systems have expanded the availability of this evidence at the clinical level, but their real-world impact depends on whether clinicians incorporate them into practice [1]. Query-log analysis has emerged as a behavioral method for characterizing the clinical information needs physicians bring to these platforms, revealing that clinical search behavior is specialized, iterative, and structurally distinct from general-purpose web search [2,3].

In Latin American health systems, the availability of digital clinical decision support tools has grown in recent years, but real-world characterizations of physician usage behavior remain limited. A systematic scoping review of artificial intelligence in low- and middle-income health systems identified only 10 real-world implementation studies and concluded that additional evidence on acceptance, reliability, and context-specific value is needed [4]. More recent reviews of large language models in clinical workflows similarly report that implementation evidence from non-high-income settings is sparse [5]. At the query level, recent studies have documented how physicians use general-purpose large language models in China [6] and the United States [7]; however, neither study examined specialized semantic clinical search tools or Latin American cohorts.

Few published descriptions exist of real-world physician query behavior in AI-enabled semantic clinical search platforms across Latin American countries. The present study aimed to characterize query topic patterns, geographic distribution, and temporal trends among Latin American physicians registered on a semantic clinical search platform during a 13-month observational period. The contribution is behavioral and descriptive: this paper documents what clinical questions Latin American physicians bring to a semantic search tool, in what proportions across countries, and how use evolved over time. It does not evaluate clinical accuracy, patient outcomes, or workflow effectiveness.

## Methods

### 2.1 Study design

We conducted a retrospective descriptive query-log analysis of anonymized physician interactions with a semantic clinical search platform deployed in Latin America. The unit of analysis varied by outcome: platform-level exports for cohort totals, country-level aggregates for geographic comparisons, monthly aggregates for temporal trends, and the full topic-field extract for category analysis. The study was reported in alignment with the STROBE statement for observational studies [8].

### 2.2 Data source and study period

Data were extracted from Arkangel platform analytics on June 17, 2026. The study period extended from May 6, 2025 (earliest query recorded in the analytics database) to June 16, 2026, covering approximately 13 months. The platform is a semantic clinical search tool developed by Arkangel AI for clinical decision support. Physicians submit natural-language queries, which the platform processes to retrieve relevant clinical evidence [9]; each query is classified by topic by the platform’s NLP layer and stored with a timestamp, anonymized user identifier, country, and the NLP-assigned topic field (queries.topic) analyzed in this study.

The first recorded query occurred between May 6 and May 18, 2025, across all 17 countries included in country-level analyses, representing a 12-day interval within the 407-day study period (2.9% of total observation time). Given this minimal variation in country entry dates, country-level comparisons were based on queries per active physician over the study period. Country start dates are reported in Table 1. Analytical files were committed to the study repository as anonymized aggregate and country-level exports. Raw query text was not retained in the study repository; a restricted de-identified sample was accessed for taxonomy validation purposes only (see Section 2.4).

**Table 1.**
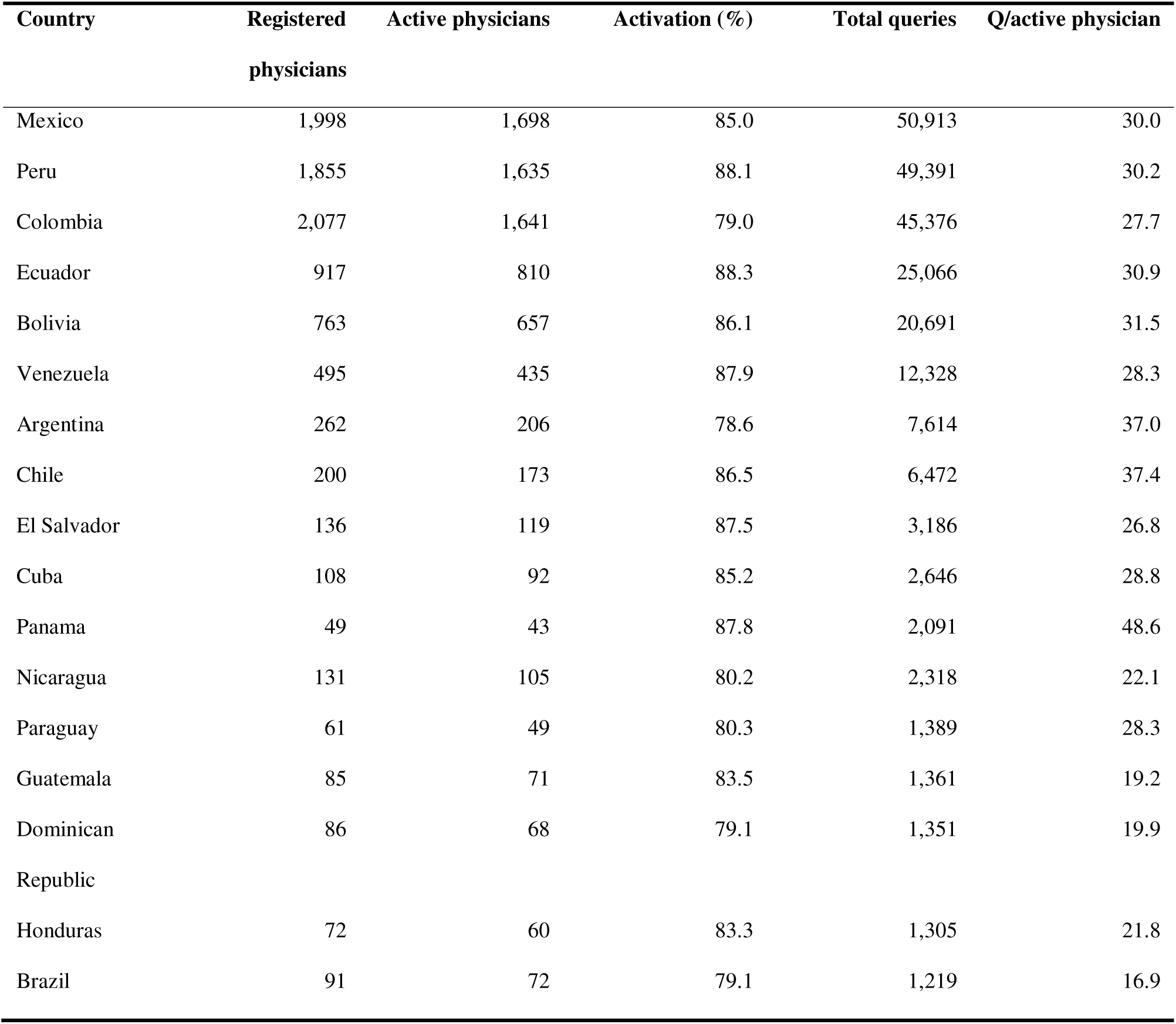
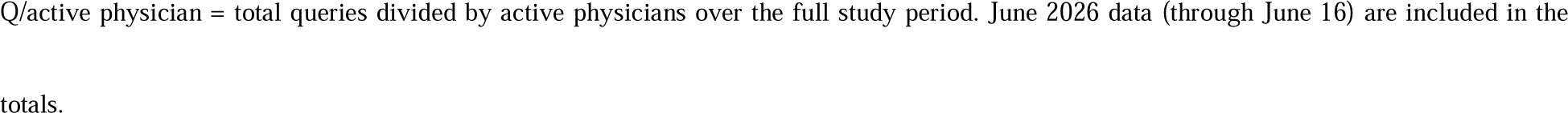
Clinical cohort, activation, queries, and usage intensity by country (17 countries with ≥1,000 queries).

### 2.3 Clinical cohort definition

The platform’s occupation field contained 619 distinct values across 109,996 registered users globally, including NULL entries (57.5%), student designations (26.3%), and other healthcare roles (6.2%). We defined the analytical cohort to include only users in clinical physician roles, using a programmatic classification of all 619 values into three groups: (a) physician core (Physician and orthographic variants, 9,116 users), (b) trainees in clinical roles (Trainee, intern, resident, and similar, 344 users), and (c) named clinical specialties with independent prescribing roles (Obstetrician, Surgeon, Radiologist, Medical Director, and 25 additional specific designations, 617 users). Medical students, users with NULL occupation fields, and other non-clinical roles were excluded from the analytical cohort. The total cohort comprised 10,077 users globally, representing 9.2% of all platform users. Of these, 9,443 were registered in Latin American countries and form the analytical sample for this study.

### 2.4 Topic taxonomy normalization

The platform queries.topic field contained 4,264 distinct raw values across Latin American queries. These values were assigned by the platform’s NLP layer at the moment of query submission and were not entered by the physician; they therefore reflect the system’s classification of each query, not a direct physician-selected label. Values showed substantial variation in phrasing, language, and specificity between countries (for example, Mexico contributed 962 distinct topic strings, of which 156 overlapped exactly with Colombian entries). The full classification pipeline is illustrated in Figure 1.

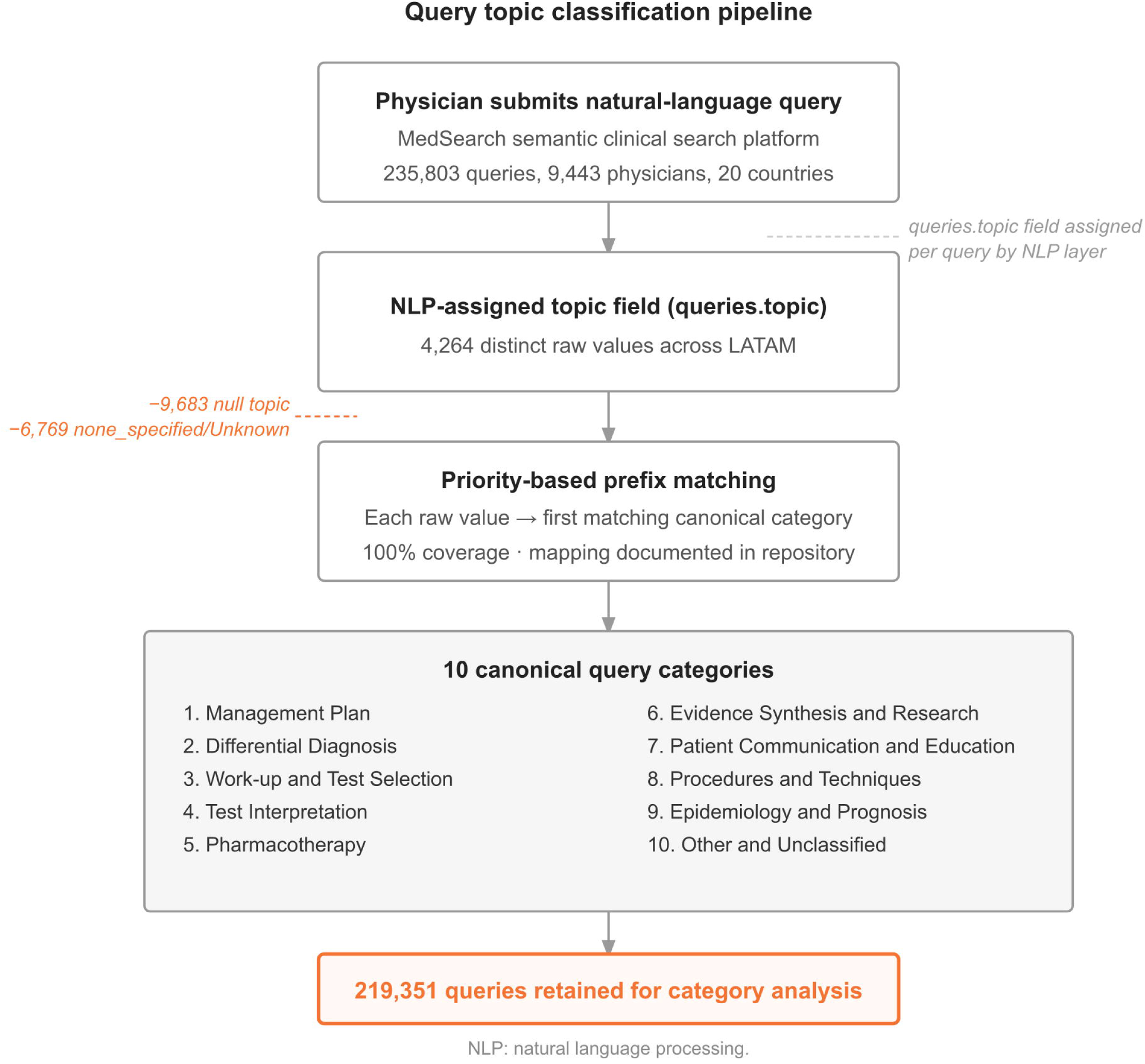

A 10-category canonical taxonomy was developed through iterative review of the most frequent values and refined iteratively until 100% coverage of all categorized queries was achieved. The taxonomy was applied using priority-based prefix matching, assigning each raw value to the first matching canonical category. Values explicitly designated as none_specified or Unknown were retained separately and excluded from the proportional topic analysis. The 10 canonical categories were Management Plan, Differential Diagnosis, Work-up and Test Selection, Test Interpretation, Pharmacotherapy, Evidence Synthesis and Research, Patient Communication and Education, Procedures and Techniques, Epidemiology and Prognosis, and Other and Unclassified.

The full classification logic is available in the study repository as Supplementary Table S1. To assess face validity of the canonical mapping, a medical epidemiologist from the study team reviewed a stratified random sample of 200 de-identified query interactions. For each sampled query, the reviewer examined both the raw query content and the corresponding NLP-assigned canonical category and assessed whether the classification was clinically appropriate. Agreement between the reviewer’s assessment and the NLP-assigned canonical category was confirmed in approximately 93% of cases. Disagreements were adjudicated through discussion with the analytic team and used to refine prefix rules before final application.

### 2.5 Variables

The primary descriptive variables were total registered physicians, activated physicians (those with at least one query), activation rate, total queries, queries per active physician over the study period, and country-level distributions of the 10 topic categories. For country-level intensity comparisons, the denominator was the number of physicians who generated at least one query (active physicians) rather than all registered physicians, to avoid diluting intensity estimates with inactive accounts. Monthly variables for the six high-volume countries (Mexico, Peru, Colombia, Ecuador, Bolivia, Venezuela) included total monthly queries. High-volume countries were defined as those with more than 10,000 queries over the study period. Monthly active physician counts by country are available in the Supplementary Table S2.

### 2.6 Privacy, anonymization, and governance

The analysis used anonymized aggregate and country-level exports. No patient names, physician names, raw query strings, or identifiable institution names appear in any analytical file. Physician identifiers were replaced with anonymized hashes in row-level files. The analysis was conducted by Arkangel AI employees on data from Arkangel’s own platform, processed under the data controller’s authorization for internal research. The dataset is compatible with Colombian Law 1581 of 2012, which governs personal data protection and authorizes data-controller processing of de-identified aggregate data for internal research purposes [10]. No patient data of any kind were used. The geographic scope of the analysis covers multiple Latin American regulatory jurisdictions; the analysis covered only aggregate behavioral data without individual patient or physician identifiers, which reduces the data protection risk profile across jurisdictions. The platform operates under internal data security and privacy controls aligned with ISO 27001:2022 and HIPAA-related safeguards, which governed the query data used in this study.

### 2.7 Analysis

We summarized categorical variables using counts and percentages. The cohort-level activation rate was reported with Wilson 95% confidence intervals. Topic category proportions were calculated among queries with a valid topic assignment (excluding null, none_specified, and Unknown values). Country-level distributions were expressed as the percentage of categorized queries assigned to each canonical topic category.

Temporal trends were examined descriptively for the six high-volume countries from May 2025 through June 2026. June 2026 represents a partial observation period (through June 16, 2026) and is presented as such.

Confidence intervals were not reported for topic category proportions because the large number of categorized queries (n=219,351) yielded intervals within approximately ±0.2 percentage points of the observed estimates, providing negligible additional interpretive value.

## Results

### 3.1 Geographic coverage and cohort

A total of 9,443 physicians across 20 Latin American countries were registered on the platform during the study period. Of these, 7,974 made at least one query (84.4%; 95% CI 83.7–85.2%), generating 235,803 queries. Seventeen countries contributed ≥1,000 queries each and were included in country-level analyses (Table 1). Three countries were below this threshold (Costa Rica, 826; Uruguay, 259; Puerto Rico, 1) and are not reported individually.

The six high-volume countries (Mexico, Peru, Colombia, Ecuador, Bolivia, Venezuela) accounted for 203,765 queries (86.4% of all LATAM queries). Per-physician query volume was broadly similar across the six high-volume countries, ranging from 27.7 (Colombia) to 31.5 (Bolivia) queries per active physician over the study period, closely clustered around the LATAM average of 29.6. Across all 17 countries the range was 16.9 (Brazil) to 48.6 (Panama, n=43 active physicians) (Table 1); the highest value was observed in Panama, although interpretation is limited by the small active cohort (n=43).

### 3.2 Query topic categories

Of 235,803 total queries, 226,120 (95.9%) had a non-null topic field. After excluding 6,769 entries designated as none_specified or Unknown, 219,351 queries were retained for category analysis. Results are summarized in Table 2.

**Table 2.**
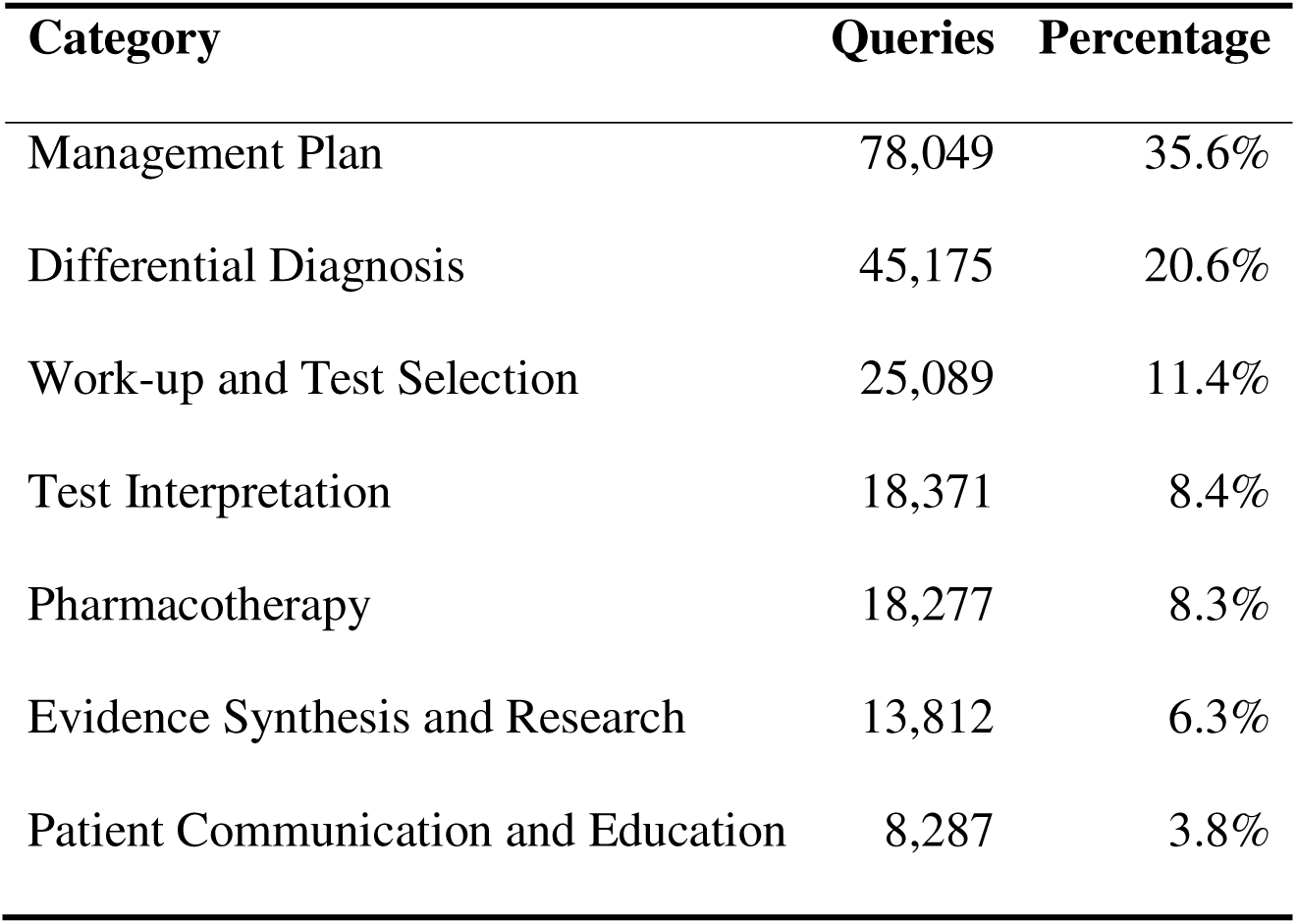
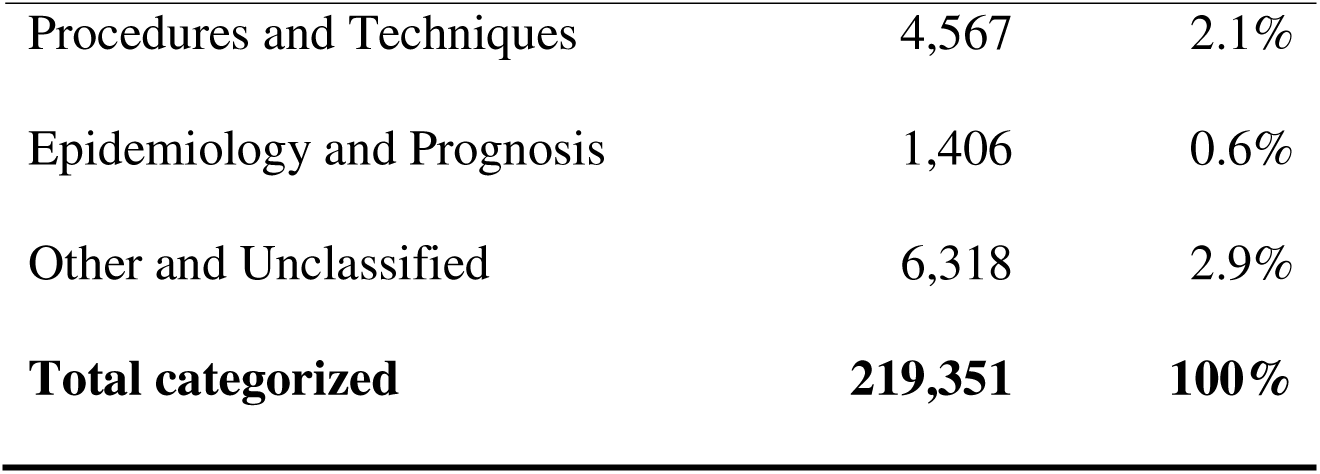
Distribution of query topic categories across 17 Latin American countries.

Management Plan was the most frequent category (78,049 queries; 35.6%), followed by Differential Diagnosis (45,175; 20.6%) and Work-up and Test Selection (25,089; 11.4%). Together, these three categories accounted for 67.6% of categorized queries. Test Interpretation (18,371; 8.4%) and Pharmacotherapy (18,277; 8.3%) each accounted for approximately 8% of queries. Evidence Synthesis and Research queries represented 6.3% of the total. Procedures and Techniques (2.1%) and Epidemiology and Prognosis (0.6%) were the least frequent categories with resolved clinical interpretations.

### 3.3 Country-level topic distribution

Management Plan was the most frequent query category in all 17 countries individually, with a range from 28.2% (Argentina) to 44.3% (Honduras). In all countries except Bolivia, the gap between Management Plan and the second-most-frequent category was at least 8 percentage points. Bolivia showed the narrowest gap, with Management Plan at 28.9% and Differential Diagnosis at 28.3%.

Test Interpretation was disproportionately frequent in six countries: Argentina (17.3%), Paraguay (15.0%), Colombia (11.4%), Brazil (10.8%), El Salvador (10.2%), and Venezuela (10.1%), compared with a global average of 8.4%. Epidemiology and Prognosis queries were below 1.5% in all 17 countries. Full country-level category distributions are presented in Table 3.

**Table 3.**
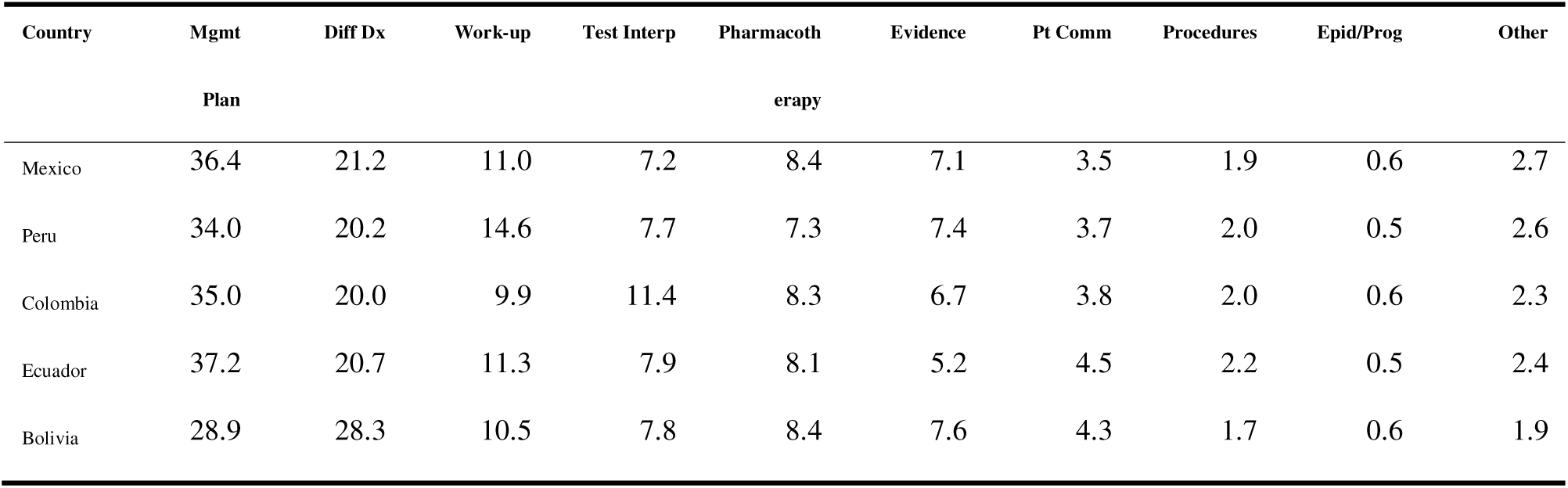
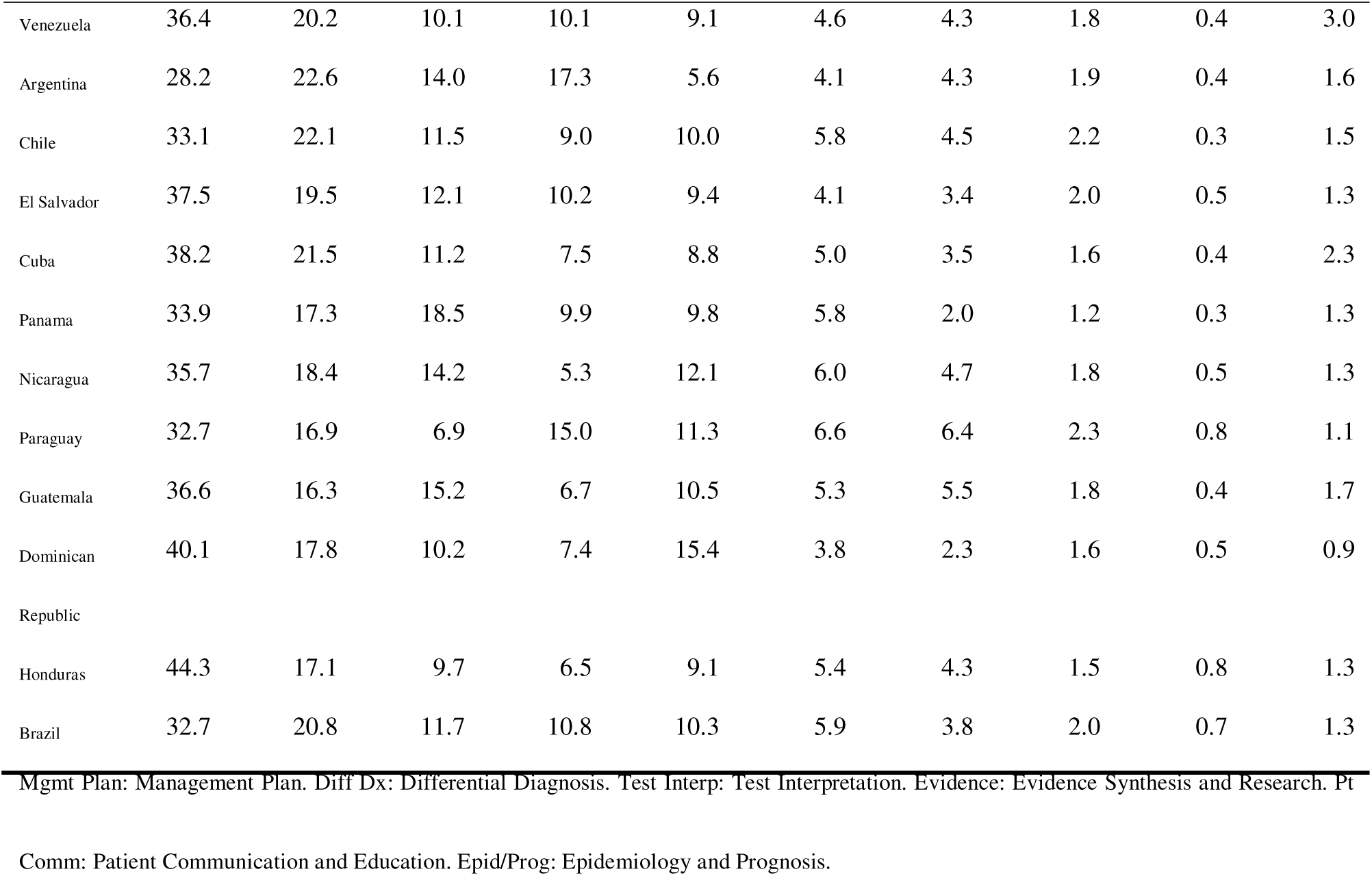
Query topic category distribution by country (proportion of topic-present queries, %). Countries ordered by total query volume.

### 3.4 Temporal trends in the six high-volume countries

Monthly query volume in the six high-volume countries (Mexico, Peru, Colombia, Ecuador, Bolivia, Venezuela) peaked in July 2025 at 29,164 queries. Country-level peaks were staggered: July 2025 for Mexico, Ecuador, Bolivia, and Venezuela; August 2025 for Colombia; and September 2025 for Peru. A sustained decline in monthly query volume was observed across all six countries from October 2025 onward, with a pronounced drop between February and March 2026 (from 12,742 to 5,469 queries, a 57.1% reduction). This concurrent pattern across geographically distinct countries is inconsistent with a purely country-specific explanation. Monthly trends are presented in Table 4.

**Table 4.** Monthly query volume for the six high-volume countries (May 2025 to June 2026).

| Month | Mexico | Peru | Colombia | Ecuador | Bolivia | Venezuela | Total 6 |
| --- | --- | --- | --- | --- | --- | --- | --- |
| May 2025 | 1,882 | 1,246 | 3,912 | 990 | 1,291 | 442 | 9,763 |
| Jun 2025 | 5,662 | 3,169 | 5,643 | 2,391 | 2,666 | 1,500 | 21,031 |
| Jul 2025 | 8,275 | 5,818 | 6,376 | 3,932 | 2,829 | 1,934 | 29,164 |
| Aug 2025 | 7,018 | 5,788 | 7,385 | 2,628 | 2,018 | 1,512 | 26,349 |
| Sep 2025 | 5,680 | 7,209 | 4,867 | 2,032 | 2,264 | 1,188 | 23,240 |
| Oct 2025 | 4,487 | 4,475 | 3,353 | 1,968 | 1,566 | 990 | 16,839 |
| Nov 2025 | 4,135 | 5,427 | 3,491 | 3,102 | 2,264 | 960 | 19,379 |
| Dec 2025 | 2,913 | 4,209 | 2,395 | 1,851 | 1,481 | 735 | 13,584 |
| Jan 2026 | 3,118 | 3,653 | 2,215 | 2,202 | 1,024 | 1,033 | 13,245 |
| Feb 2026 | 3,295 | 3,106 | 2,430 | 1,848 | 1,121 | 942 | 12,742 |
| Mar 2026 | 1,380 | 1,314 | 1,115 | 756 | 536 | 368 | 5,469 |
| Apr 2026 | 1,329 | 1,559 | 1,025 | 605 | 606 | 278 | 5,402 |
| May 2026 | 1,238 | 1,498 | 758 | 491 | 713 | 236 | 4,934 |
| Jun 2026* | 730 | 937 | 414 | 272 | 319 | 210 | 2,882 |

\*June 2026 is a partial period (data through June 16, 2026 only).

## Discussion

### 4.1 Principal findings

In this retrospective query-log analysis of 9,443 Latin American physicians, clinical search was oriented predominantly toward clinical decision support: Management Plan, Differential Diagnosis, and Work-up and Test Selection together accounted for 67.6% of categorized queries. Management Plan was the most frequent category in all 17 countries individually, with a range of 28.2% to 44.3%. This cross-country consistency is a central finding of this study. Regardless of country size, physician cohort, or usage intensity, the hierarchy of clinical information needs showed the same top category, suggesting that therapeutic planning represents a relatively consistent demand signal across Latin American countries.

Per-physician query volume was broadly similar across the six high-volume countries (27.7 to 31.5 queries per active physician over the study period, closely clustered around the LATAM average of 29.6), indicating that differences in total query volume reflected primarily variation in enrolled cohort size. Countries with larger physician cohorts generated more total queries, but at similar rates per active physician. This consistency suggests that physicians who activated the platform showed similar search intensity across countries.

The concurrent decline in monthly query volume across all six high-volume countries from October 2025 onward, including a 57.1% reduction between February and March 2026, suggests that the pattern was not confined to a single national context. The decline occurred simultaneously across Mexico, Peru, Colombia, Ecuador, Bolivia, and Venezuela despite substantial differences in cohort size and healthcare settings. Whether this pattern reflects platform-level factors, shared regional academic or calendar cycles, or other external influences cannot be determined from the available data.

Prior query-log studies of clinical information behavior provide context for these findings. Yang and colleagues, analyzing 202,905 queries from 533 users in an electronic health record search engine, found that clinical queries were specialized, iterative, and terminologically distinct from web search [2]. Natarajan and colleagues similarly documented clinical information needs through query-log analysis in an EHR search utility and found that diagnostic and management queries predominated [3]. The distribution observed here, with management and diagnostic queries together exceeding 56% of categorized volume, is broadly consistent with those findings from different settings, suggesting some cross-system stability in clinical information needs at the point of care. Prospective and mixed-methods studies of AI-enabled clinical decision support in primary care have found that physicians generally perceive these systems as trustworthy and easy to use, while noting difficulty in establishing clinical outcomes impact [11,12]. Systematic reviews of clinical AI acceptance have documented variable adoption across settings, with workflow fit and trustworthiness as consistent determinants [13]. This distinction applies equally here: query volume and category distribution describe information-seeking behavior, not decision quality or clinical effectiveness.

Unlike the earlier studies, which captured queries within a single institutional EHR system in the United States, the present analysis covers 17 countries and reflects access to a dedicated semantic clinical search platform rather than a general-purpose EHR search utility. The cross-country consistency in category distribution, despite differences in national health system structure, specialty density, and institutional context, is a finding not previously documented for Latin American settings. Test Interpretation showed a notable cluster of countries with elevated frequency (Argentina, Paraguay, Colombia, Brazil, El Salvador, Venezuela all above 10%) compared with a global average of 8.4%. Whether this pattern reflects practice context, specialty distribution, or query formulation habits in those cohorts is not answerable from the current data. Proportional distributions in the nine countries with fewer than 120 active physicians (El Salvador, Cuba, Panama, Nicaragua, Paraguay, Guatemala, Dominican Republic, Honduras, Brazil) warrant cautious interpretation, as small cohort sizes increase the influence of individual usage patterns on country-level percentages.

Epidemiology and Prognosis queries were below 1.5% in all countries. This finding is consistent with the clinical positioning of the tool as a decision support resource rather than an epidemiological reference: physicians using the platform to support individual patient decisions are more likely to submit queries about what to do or what to investigate than about population-level disease incidence or prognosis distributions.

One alternative interpretation of the category distribution is that it reflects the platform’s design rather than physicians’ underlying information needs: if the tool’s query architecture was built around management planning and diagnostic categories, finding those categories dominant could be a design artifact. Several observations suggest that platform design is unlikely to fully explain the observed distribution. First, category distributions varied meaningfully across countries: the range for Management Plan alone was 28.2% to 44.3%, a 16-point spread that would not be expected if platform design fully determined the distribution. Second, the elevated Test Interpretation frequency observed in six countries (Argentina, Paraguay, Colombia, Brazil, El Salvador, Venezuela) was not a structural feature of the platform’s original design emphasis. Third, the dominance of management and diagnostic queries is consistent with findings from EHR query-log studies conducted in different systems and countries [2,3], which were not built on the same platform. Together, these observations support the interpretation that the category hierarchy reflects genuine clinical information needs, while acknowledging that platform design may exert some influence that cannot be fully disentangled from a single-platform dataset.

### 4.2 Implications for Latin American implementation

While the broader literature on AI in clinical medicine has grown substantially [14,15], evidence on AI-enabled clinical decision support in low- and middle-income health systems remains limited. Ciecierski-Holmes and colleagues identified only 10 real-world LMIC AI health studies and highlighted barriers including data availability, trust, interpretability, and cost-effectiveness [4]. Recent reviews of large language models in clinical workflows similarly emphasize that implementation evidence from non-high-income settings is sparse [5]. The behavioral profile documented here, where physicians across 17 countries consistently directed their queries toward management planning, differential diagnosis, and test selection, provides a descriptive baseline for designing tools and evaluating whether semantic clinical search addresses the information needs physicians actually bring to clinical encounters. The broader clinical decision support literature indicates that successful deployment depends on workflow integration, usability, and governance rather than model performance alone [16–20]. For AI-enabled tools specifically, implementation evidence also depends on explainability, clinician trust, and human-centered evaluation [21–25], factors particularly relevant in settings where digital health governance frameworks are still developing.

The uniformity in per-physician query volume across countries (16.9 to 48.6 queries per active physician, median approximately 29) indicates that physicians who activate the platform use it at broadly similar rates. The primary driver of total query volume differences between countries was cohort size, not usage intensity per physician. Future studies should evaluate how information-seeking patterns relate to physician performance, workflow outcomes, and clinical decision-making.

The concurrent regional decline in query volume from October 2025 onward raises a governance question relevant to all multi-country digital health deployments: what monitoring mechanisms would detect a simultaneous usage decline across countries in time to investigate and respond? The present analysis identified the pattern retrospectively. Prospective dashboard monitoring of monthly active physicians by country would provide earlier visibility into regional trends and allow platform operators to distinguish cohort attrition from access disruptions.

### 4.3 Strengths and limitations

This study has several methodological strengths. The cohort spans 20 Latin American countries over 13 months, making it one of the largest multi-country behavioral analyses of clinical search use published from the region. The analysis is based on passive platform activity rather than self-reported survey data, which eliminates recall bias and response-set effects. Coverage within the enrolled cohort is complete: all physician queries generated during the study period were included, not a sample. The topic taxonomy achieved 100% coverage of 4,264 distinct NLP-assigned values, and the analytical pipeline is fully documented and reproducible.

Several limitations should be considered alongside these strengths. The analytical cohort was defined using the platform’s occupation field, which was NULL for 57.5% of all registered users; clinical physicians who registered with atypical labels or no occupation entry are not captured, and the cohort likely undercounts total physician activity on the platform. Additionally, the topic taxonomy was derived from the platform’s queries.topic field, which reflects the NLP layer’s classification of each query rather than a physician-selected label, and the 10 categories were not validated against an external taxonomy (for example, MeSH clinical query filters) or through inter-rater reliability assessment; to mitigate this, internal consistency checks were applied, the hierarchy was consistent across 17 independent country cohorts, and a medical epidemiologist from the study team reviewed a stratified random sample of 200 query interactions, examining both the raw query content and the NLP-assigned canonical category, confirming agreement in approximately 93% of cases (see Section 2.4).

A further limitation is that the study is observational and descriptive, without a control group, physician satisfaction data, or patient outcome information; query volume and category distribution describe information-seeking behavior, not clinical effectiveness. Activation rates and query intensity may also partly reflect differences in platform deployment and onboarding by country, which the current data do not allow separation of. Finally, the data come from a single platform operated by a single vendor, which limits generalizability to other clinical search tools or Latin American health systems. The concurrent regional decline in query volume from October 2025 onward could not be attributed to a specific cause from the available data.

### 4.4 Future research

Future studies should evaluate whether semantic clinical search improves time to answer, perceived usefulness, physician confidence, or guideline-concordant decisions, combining usage logs with prospective physician surveys and task-based evaluations. Prospective studies examining whether activation rates correlate with structured onboarding programs are warranted, using the baseline documented here as a comparator. Validation of the topic taxonomy against manually annotated queries and inter-rater reliability assessment would strengthen confidence in the category structure before clinical inferences are made. Studies evaluating the concurrent regional decline through real-time monitoring dashboards combined with institutional access records would clarify whether the pattern reflects cohort attrition, platform changes, or calendar effects.

## Conclusion

In this multi-country analysis of 219,351 clinical queries generated by physicians across 20 Latin American countries, information seeking was dominated by management planning, diagnostic reasoning, and test selection. Topic distributions were broadly consistent across countries despite differences in cohort size and healthcare settings. These findings provide a regional behavioral baseline for understanding physician information needs in semantic clinical search systems.

## Data availability

Anonymized aggregate and country-level data files used in this analysis are available from the corresponding author upon reasonable request. The files are held in Arkangel AI’s internal research repository and are not publicly accessible due to platform data governance requirements. Raw query text was not retained for analysis and is not available for release.

## Ethics and privacy statement

This study involved retrospective analysis of anonymized aggregate usage logs from Arkangel AI’s own platform. No patient data, patient identifiers, physician names, raw query text, or identifiable institution names were used at any stage of the analysis. All data were de-identified country-level aggregates processed entirely by the data controller (Arkangel AI) under explicit authorization from company leadership.

Under Colombian Ministerio de Salud Resolución 8430 de 1993, which establishes the scientific, technical, and administrative standards for health research in Colombia, this study is classified as *investigación sin riesgo* (no-risk research): it involves retrospective documentary analysis of anonymized records with no intervention or modification of any biological, physiological, psychological, or social variable of any participant [26]. Accordingly, external ethics committee review was not required. The study is also compatible with Colombian Ley 1581 de 2012, which authorizes data controllers to process de-identified data for internal research purposes [10]. The analysis was conducted in accordance with the general ethical principles for health-related research set out in the World Medical Association Declaration of Helsinki [27] and the CIOMS International Ethical Guidelines for Health-related Research Involving Humans [28].

## Acknowledgments

The authors thank the physicians across Latin America who used the Arkangel platform (formerly MedSearch) and whose clinical queries made this analysis posible.

Portions of this manuscript were drafted and revised with the assistance of Gabo, Arkangel AI’s internal research writing agent (developed using OpenAI Codex), and Claude (Anthropic), both used as writing aids under human supervision. All content, analyses, interpretations, and scientific judgments were reviewed, verified, and approved by the human authors.

## References

1. Khairat S, Marc D, Crosby W, Al Sanousi A. Reasons for physicians not adopting clinical decision support systems: critical analysis. JMIR Med Inform. 2018;6(2):e24. doi:10.2196/medinform.8912

2. Yang L, Mei Q, Zheng K, Hanauer DA. Query log analysis of an electronic health record search engine. AMIA Annu Symp Proc. 2011;2011:915–924. PMID:22195150.

3. Natarajan K, Stein D, Jain S, Elhadad N. An analysis of clinical queries in an electronic health record search utility. Int J Med Inform. 2010;79(7):515–522. doi:10.1016/j.ijmedinf.2010.03.004

4. Ciecierski-Holmes T, Singh R, Axt M, Brenner S, Barteit S. Artificial intelligence for strengthening healthcare systems in low- and middle-income countries: a systematic scoping review. NPJ Digit Med. 2022;5:162. doi:10.1038/s41746-022-00700-y

5. Artsi Y, Sorin V, Glicksberg BS, Korfiatis P, Nadkarni GN, Klang E. Large language models in real-world clinical workflows: a systematic review of applications and implementation. Front Digit Health. 2025;7:1659134. doi:10.3389/fdgth.2025.1659134

6. Qiu L, Tang C, Bi X, Burtch G, Chen Y, Zhang H. Physician use of large language models: a quantitative study based on large-scale query-level data. J Med Internet Res. 2025;27:e76941. doi:10.2196/76941

7. Unell A, Kashyap M, Pfeffer M, Shah N, et al. Real-world usage patterns of large language models in healthcare. medRxiv. 2025. doi:10.1101/2025.05.02.25326781

8. von Elm E, Altman DG, Egger M, Pocock SJ, Gøtzsche P, Vandenbroucke JP, et al. The Strengthening the Reporting of Observational Studies in Epidemiology (STROBE) statement: guidelines for reporting observational studies. PLoS Med. 2007;4(10):e296. doi:10.1371/journal.pmed.0040296

9. Villa MC, Castaño-Villegas N, Llano I, Martinez J, Guevara MF, Zea J, Velásquez L. Arkangel AI: a conversational agent for real-time, evidence-based medical question-answering. Intell Based Med. 2025;12:100274. doi:10.1016/j.ibmed.2025.100274

10. Congreso de la República de Colombia. Ley Estatutaria 1581 de 2012. Diario Oficial No. 48.587. 2012. URL: https://observatoriolegislativocele.com/en/colombia-personal-data-protection-law-2012

11. Herter WE, Khuc J, Bonten TN, Verheij RA, Numans ME, Chavannes NH. Usability and usefulness of machine learning-based clinical decision support software in primary care: survey of users in a prospective observational study. JMIR Med Inform. 2026;14:e80527. doi:10.2196/80527

12. Allen MR, Webb S, Mandvi A, Frieden M, Tai-Seale M, Kallenberg G. Navigating the doctor-patient-AI relationship: a mixed-methods study of physician attitudes toward artificial intelligence in primary care. BMC Prim Care. 2024;25(1):42. doi:10.1186/s12875-024-02282-y

13. Chen M, Zhang B, Cai Z, Seery S, Gonzalez MJ, Ali NM, Ren R, Qiao Y, Xue P, Jiang Y. Acceptance of clinical artificial intelligence among physicians and medical students: a systematic review with cross-sectional survey. Front Med (Lausanne). 2022;9:990604. doi:10.3389/fmed.2022.990604

14. Topol EJ. High-performance medicine: the convergence of human and artificial intelligence. Nat Med. 2019;25(1):44–56. doi:10.1038/s41591-018-0300-7

15. Rajkomar A, Dean J, Kohane I. Machine learning in medicine. N Engl J Med. 2019;380(14):1347–1358. doi:10.1056/NEJMra1814259

16. Sutton RT, Pincock D, Baumgart DC, Sadowski DC, Fedorak RN, Kroeker KI. An overview of clinical decision support systems: benefits, risks, and strategies for success. NPJ Digit Med. 2020;3:17. doi:10.1038/s41746-020-0221-y

17. Kawamoto K, Houlihan CA, Balas EA, Lobach DF. Improving clinical practice using clinical decision support systems: a systematic review of trials to identify features critical to success. BMJ. 2005;330(7494):765. doi:10.1136/bmj.38398.500764.8F

18. Bates DW, Kuperman GJ, Wang S, Gandhi T, Kittler A, Volk L, et al. Ten Commandments for Effective Clinical Decision Support: Making the Practice of Evidence-based Medicine a Reality. J Am Med Inform Assoc. 2003;10(6):523–530. doi:10.1197/jamia.M1370

19. Bright TJ, Wong A, Dhurjati R, Bristow E, Bastian L, Coeytaux RR, et al. Effect of clinical decision-support systems: a systematic review. Ann Intern Med. 2012;157(1):29–43. doi:10.7326/0003-4819-157-1-201207030-00450

20. Sittig DF, Wright A, Osheroff JA, Middleton B, Teich JM, Ash JS, et al. Grand challenges in clinical decision support. J Biomed Inform. 2008;41(2):387–392. doi:10.1016/j.jbi.2007.09.003

21. Kelly CJ, Karthikesalingam A, Suleyman M, Corrado G, King D. Key challenges for delivering clinical impact with artificial intelligence. BMC Med. 2019;17:195. doi:10.1186/s12916-019-1426-2

22. Wiens J, Saria S, Sendak M, Ghassemi M, Liu VX, Doshi-Velez F, et al. Do no harm: a roadmap for responsible machine learning for health care. Nat Med. 2019;25(9):1337–1340. doi:10.1038/s41591-019-0548-6

23. Amann J, Blasimme A, Vayena E, Frey D, Madai VI. Explainability for artificial intelligence in healthcare: a multidisciplinary perspective. BMC Med Inform Decis Mak. 2020;20:310. doi:10.1186/s12911-020-01332-6

24. Beede E, Baylor E, Hersch F, Iurchenko A, Wilcox L, Ruamviboonsuk P, et al. A human-centered evaluation of a deep learning system deployed in clinics for the detection of diabetic retinopathy. Proc CHI Conf Hum Factors Comput Syst. 2020:1–12. doi:10.1145/3313831.3376718

25. Wang D, Zhang Y, Wang Y, Cao J, Wang Q, Bian J, et al. Human-centered design and evaluation of AI-empowered clinical decision support systems: a systematic review. Front Comput Sci. 2023;5:1187299. doi:10.3389/fcomp.2023.1187299

26. Colombia. Ministerio de Salud. Resolución 8430 de 1993: Por la cual se establecen las normas científicas, técnicas y administrativas para la investigación en salud. Bogotá: Ministerio de Salud; 1993. URL: https://www.minsalud.gov.co/sites/rid/Lists/BibliotecaDigital/RIDE/DE/DIJ/RESOLUCION-8430-DE-1993.PDF

27. World Medical Association. Declaration of Helsinki: Ethical Principles for Medical Research Involving Human Subjects. JAMA. 2013;310(20):2191–2194. doi:10.1001/jama.2013.281053

28. Council for International Organizations of Medical Sciences (CIOMS). International Ethical Guidelines for Health-related Research Involving Humans. Geneva: CIOMS; 2016. URL: https://cioms.ch/publications/product/international-ethical-guidelines-for-health-related-research-involving-humans/

