## Supplementary-material for "Clinical Information Needs Among Latin American Physicians: A Multi-Country Analysis of Semantic Clinical Search"

**Supplementary Table S1. Taxonomy Validation — Face validity review of 200 sampled query classifications.**

| **Sample ID** | **Country** | **Query Date** | **NLP-Assigned Topic** | **Reviewer Category** | **Agreement** | **Reviewer Note** |
| --- | --- | --- | --- | --- | --- | --- |
| 1 | Perú | 2025-08-20 | Prevention & Screening: Primary/secondary prevention | Prevention & Screening: Primary/secondary prevention | Match | OK |
| 2 | Ecuador | 2025-10-15 | Patient Communication: Translate to lay language | Patient Communication: Translate to lay language | Match | OK |
| 3 | Venezuela | 2025-08-24 | Test Interpretation | Test Interpretation | Match | OK |
| 4 | México | 2026-01-15 | Work-up & Test Selection: Decide which tests/imaging to order | Work-up & Test Selection: Decide which tests/imaging to order | Match | OK |
| 5 | Argentina | 2025-05-12 | Differential Diagnosis | Differential Diagnosis | Match | OK |
| 6 | México | 2025-11-19 | Differential Diagnosis: Identify likely conditions explaining a presentation | Differential Diagnosis: Identify likely conditions explaining a presentation | Match | OK |
| 7 | México | 2025-11-12 | Management Plan: Choose therapeutic strategy (non-drug + drug) | Management Plan: Choose therapeutic strategy (non-drug + drug) | Match | OK |
| 8 | México | 2025-10-16 | Test Interpretation: Interpret numbers, visuals or cut-offs | Test Interpretation: Interpret numbers, visuals or cut-offs | Match | OK |
| 9 | México | 2025-07-15 | Differential Diagnosis | Differential Diagnosis | Match | OK |
| 10 | Perú | 2025-06-17 | Evidence Synthesis / Research | Evidence Synthesis / Research | Match | OK |
| 11 | Ecuador | 2026-03-13 | Patient Communication: Translate to lay language | Evidence Synthesis / Research | Review | CHANGE: Classified as Patient Communication but content consists of bibliographic resources for teaching costal anatomy, not patient-facing translation |
| 12 | Venezuela | 2025-07-04 | Procedures & Techniques | Procedures & Techniques | Match | OK |
| 13 | Colombia | 2025-11-14 | Management Plan: Choose therapeutic strategy (non-drug + drug) | Management Plan: Choose therapeutic strategy (non-drug + drug) | Match | OK |
| 14 | Bolivia | 2025-06-07 | Summary / Overview | Summary / Overview | Match | OK |
| 15 | Ecuador | 2025-09-26 | Mechanisms of Physical and Chemical Defense | Evidence Synthesis / Research | Review | CHANGE: Non-canonical label ('Mechanisms of Physical and Chemical Defense'); content is a synthesis of basic physiology |
| 16 | Perú | 2025-10-03 | Guideline Lookup | Guideline Lookup | Match | OK |
| 17 | Chile | 2026-03-13 | Management Plan: Choose therapeutic strategy (non-drug + drug) | Management Plan: Choose therapeutic strategy (non-drug + drug) | Match | OK |
| 18 | Bolivia | 2025-08-08 | Evidence Synthesis / Research | Evidence Synthesis / Research | Match | OK |
| 19 | México | 2025-06-15 | Patient Communication | Patient Communication | Match | OK |
| 20 | Perú | 2025-12-12 | Work-up & Test Selection: Decide which tests/imaging to order | Work-up & Test Selection: Decide which tests/imaging to order | Match | OK |
| 21 | Perú | 2025-10-25 | Management Plan: Choose therapeutic strategy (non-drug + drug) | Management Plan: Choose therapeutic strategy (non-drug + drug) | Match | OK |
| 22 | México | 2025-07-16 | nan | Management Plan | Review | NOT CLASSIFIED IN DATABASE: content on cervical collar management, physiotherapy, and road accident injuries |
| 23 | Bolivia | 2025-12-12 | Work-up & Test Selection: Decide which tests/imaging to order | Work-up & Test Selection: Decide which tests/imaging to order | Match | OK |
| 24 | Perú | 2025-09-09 | nan | Management Plan | Review | NOT CLASSIFIED IN DATABASE: direct query on updated treatment for gastritis |
| 25 | Argentina | 2025-06-05 | Differential Diagnosis | Differential Diagnosis | Match | OK |
| 26 | Bolivia | 2025-08-25 | Evidence Synthesis / Research | Evidence Synthesis / Research | Match | OK |
| 27 | Perú | 2025-08-11 | Work-up & Test Selection | Work-up & Test Selection | Match | OK |
| 28 | Argentina | 2026-02-25 | Prognosis & Follow-up: Expected outcomes and monitoring | Prognosis & Follow-up: Expected outcomes and monitoring | Match | OK |
| 29 | Colombia | 2026-05-12 | Management Plan: Choose therapeutic strategy (non-drug + drug) | Management Plan: Choose therapeutic strategy (non-drug + drug) | Match | OK |
| 30 | México | 2025-07-20 | Management Plan | Management Plan | Match | OK |
| 31 | Colombia | 2025-09-17 | Differential Diagnosis and Management Plan | Differential Diagnosis and Management Plan | Match | OK |
| 32 | Colombia | 2025-08-25 | Management Plan | Management Plan | Match | OK |
| 33 | Colombia | 2025-05-29 | Differential Diagnosis | Differential Diagnosis | Match | OK |
| 34 | Perú | 2025-09-09 | Evidence Synthesis / Research | Evidence Synthesis / Research | Match | OK |
| 35 | Colombia | 2025-06-05 | Management Plan | Management Plan | Match | OK |
| 36 | Argentina | 2025-08-18 | Evidence Synthesis / Research | Evidence Synthesis / Research | Match | OK |
| 37 | México | 2026-02-08 | Management Plan: Choose therapeutic strategy (non-drug + drug) | Management Plan: Choose therapeutic strategy (non-drug + drug) | Match | OK |
| 38 | Perú | 2025-09-07 | Guideline Lookup | Guideline Lookup | Match | OK |
| 39 | Perú | 2025-08-02 | Differential Diagnosis | Differential Diagnosis | Match | OK |
| 40 | Bolivia | 2025-11-07 | Differential Diagnosis: Identify likely conditions explaining a presentation | Differential Diagnosis: Identify likely conditions explaining a presentation | Match | OK |
| 41 | Bolivia | 2025-09-24 | Differential Diagnosis | Differential Diagnosis | Match | OK |
| 42 | Ecuador | 2026-06-17 | Management Plan: Choose therapeutic strategy (non-drug + drug) | Management Plan: Choose therapeutic strategy (non-drug + drug) | Match | OK |
| 43 | Colombia | 2025-07-30 | Management Plan | Management Plan | Match | OK |
| 44 | Chile | 2025-06-09 | Differential Diagnosis / Prognosis & Follow-up | Differential Diagnosis / Prognosis & Follow-up | Match | OK |
| 45 | Bolivia | 2025-11-12 | Management Plan: Choose therapeutic strategy (non-drug + drug) | Management Plan: Choose therapeutic strategy (non-drug + drug) | Match | OK |
| 46 | Venezuela | 2025-09-23 | Differential Diagnosis | Differential Diagnosis | Match | OK |
| 47 | México | 2025-07-25 | Procedures & Techniques | Procedures & Techniques | Match | OK |
| 48 | Perú | 2025-09-03 | Management Plan | Management Plan | Match | OK |
| 49 | Colombia | 2025-07-09 | Guideline Lookup | Guideline Lookup | Match | OK |
| 50 | Ecuador | 2025-12-26 | Differential Diagnosis: Identify likely conditions explaining a presentation | Differential Diagnosis: Identify likely conditions explaining a presentation | Match | OK |
| 51 | Colombia | 2025-06-21 | Management Plan | Management Plan | Match | OK |
| 52 | México | 2025-07-15 | Test Interpretation | Evidence Synthesis / Research | Review | CHANGE: Classified as Test Interpretation but concerns water determination in nutrition/metabolism, not interpretation of a specific clinical test |
| 53 | México | 2025-09-26 | Differential Diagnosis | Differential Diagnosis | Match | OK |
| 54 | México | 2026-01-14 | Differential Diagnosis: Identify likely conditions explaining a presentation | Differential Diagnosis: Identify likely conditions explaining a presentation | Match | OK |
| 55 | México | 2025-07-19 | Management Plan | Management Plan | Match | OK |
| 56 | Ecuador | 2025-05-25 | Procedures & Techniques | Procedures & Techniques | Match | OK |
| 57 | Colombia | 2025-08-21 | nan | Test Interpretation | Review | NOT CLASSIFIED IN DATABASE: direct question about radiographic findings in a child |
| 58 | Venezuela | 2026-05-24 | Guideline Lookup: Locate authoritative guidance | Guideline Lookup: Locate authoritative guidance | Match | OK |
| 59 | Colombia | 2025-07-07 | Management Plan | Management Plan | Match | OK |
| 60 | Ecuador | 2026-02-07 | Differential Diagnosis: Identify likely conditions explaining a presentation | Differential Diagnosis: Identify likely conditions explaining a presentation | Match | OK |
| 61 | México | 2025-07-18 | Management Plan | Management Plan | Match | OK |
| 62 | México | 2025-07-30 | Guideline Lookup | Guideline Lookup | Match | OK |
| 63 | Perú | 2025-09-20 | Evidence Synthesis / Research | Evidence Synthesis / Research | Match | OK |
| 64 | Perú | 2026-02-15 | Differential Diagnosis: Identify likely conditions explaining a presentation | Differential Diagnosis: Identify likely conditions explaining a presentation | Match | OK |
| 65 | Venezuela | 2026-02-05 | Differential Diagnosis: Identify likely conditions explaining a presentation | Summary / Overview | Review | CHANGE: Classified as Differential Diagnosis but is a conceptual definition of outpatient vs. inpatient care, without a clinical presentation |
| 66 | Perú | 2025-10-31 | Pharmacotherapy: Precise drug information | Pharmacotherapy: Precise drug information | Match | OK |
| 67 | El Salvador | 2025-11-04 | Differential Diagnosis: Identify likely conditions explaining a presentation | Differential Diagnosis: Identify likely conditions explaining a presentation | Match | OK |
| 68 | Perú | 2026-01-27 | Work-up & Test Selection: Decide which tests/imaging to order | Work-up & Test Selection: Decide which tests/imaging to order | Match | OK |
| 69 | Perú | 2026-04-17 | Management Plan: Choose therapeutic strategy (non-drug + drug) | Management Plan: Choose therapeutic strategy (non-drug + drug) | Match | OK |
| 70 | Perú | 2025-06-06 | Differential Diagnosis | Differential Diagnosis | Match | OK |
| 71 | Bolivia | 2026-05-05 | Management Plan: Choose therapeutic strategy (non-drug + drug) | Management Plan: Choose therapeutic strategy (non-drug + drug) | Match | OK |
| 72 | Ecuador | 2025-08-15 | nan | Work-up & Test Selection | Review | NOT CLASSIFIED IN DATABASE: requests complete diagnostic workup for suspected acute leukemia with pancytopenia |
| 73 | México | 2026-02-12 | Work-up & Test Selection: Decide which tests/imaging to order | Work-up & Test Selection: Decide which tests/imaging to order | Match | OK |
| 74 | Argentina | 2025-09-08 | Pharmacotherapy | Pharmacotherapy | Match | OK |
| 75 | México | 2026-05-23 | Management Plan: Choose therapeutic strategy (non-drug + drug) | Management Plan: Choose therapeutic strategy (non-drug + drug) | Match | OK |
| 76 | Colombia | 2026-05-22 | Management Plan: Choose therapeutic strategy (non-drug + drug) | Patient Communication | Review | NOT CLASSIFIED IN DATABASE: conceptual question about what allergic rhinorrhea is |
| 77 | México | 2025-11-11 | Test Interpretation: Interpret numbers, visuals or cut-offs | Test Interpretation: Interpret numbers, visuals or cut-offs | Match | OK |
| 78 | Bolivia | 2025-06-06 | Management Plan | Management Plan | Match | OK |
| 79 | Colombia | 2025-06-26 | Test Interpretation | Test Interpretation | Match | OK |
| 80 | Colombia | 2025-09-22 | Management Plan | Management Plan | Match | OK |
| 81 | Nicaragua | 2025-09-08 | Prognosis & Follow-up | Prognosis & Follow-up | Match | OK |
| 82 | Bolivia | 2025-07-10 | Pharmacotherapy | Pharmacotherapy | Match | OK |
| 83 | Perú | 2025-12-10 | Differential Diagnosis: Identify likely conditions explaining a presentation | Differential Diagnosis: Identify likely conditions explaining a presentation | Match | OK |
| 84 | El Salvador | 2025-07-01 | Pharmacotherapy | Pharmacotherapy | Match | OK |
| 85 | México | 2025-08-29 | Management Plan | Management Plan | Match | OK |
| 86 | Venezuela | 2025-06-01 | Hormonal Regulation and Feedback in Reproductive Physiology | Evidence Synthesis / Research | Review | CHANGE: Non-canonical label ('Hormonal Regulation and Feedback in Reproductive Physiology'); content is a synthesis of reproductive physiology |
| 87 | México | 2026-02-19 | Differential Diagnosis: Identify likely conditions explaining a presentation | Differential Diagnosis: Identify likely conditions explaining a presentation | Match | OK |
| 88 | Venezuela | 2025-05-27 | Evidence Synthesis / Research | Evidence Synthesis / Research | Match | OK |
| 89 | Perú | 2025-06-03 | General / Other | General / Other | Match | OK |
| 90 | Venezuela | 2025-08-12 | Management Plan | Management Plan | Match | OK |
| 91 | Colombia | 2025-10-22 | Test Interpretation: Interpret numbers, visuals or cut-offs | Test Interpretation: Interpret numbers, visuals or cut-offs | Match | OK |
| 92 | Colombia | 2025-08-18 | Differential Diagnosis | Differential Diagnosis | Match | OK |
| 93 | Colombia | 2026-04-29 | Guideline Lookup: Locate authoritative guidance | Guideline Lookup: Locate authoritative guidance | Match | OK |
| 94 | Perú | 2025-10-17 | Differential Diagnosis: Identify likely conditions explaining a presentation | Evidence Synthesis / Research | Review | CHANGE: Classified as Differential Diagnosis but is a review of immunology concepts for an exam; no clinical presentation |
| 95 | Venezuela | 2025-12-26 | Differential Diagnosis: Identify likely conditions explaining a presentation | Differential Diagnosis: Identify likely conditions explaining a presentation | Match | OK |
| 96 | Bolivia | 2026-02-01 | Work-up & Test Selection: Decide which tests/imaging to order | Work-up & Test Selection: Decide which tests/imaging to order | Match | OK |
| 97 | Perú | 2026-01-01 | Management Plan: Choose therapeutic strategy (non-drug + drug) | Management Plan: Choose therapeutic strategy (non-drug + drug) | Match | OK |
| 98 | Venezuela | 2025-06-16 | Differential Diagnosis | Differential Diagnosis | Match | OK |
| 99 | México | 2025-08-12 | Pharmacotherapy | Pharmacotherapy | Match | OK |
| 100 | Ecuador | 2025-05-13 | Evidence Synthesis / Research | Evidence Synthesis / Research | Match | OK |
| 101 | Ecuador | 2026-03-28 | Management Plan: Choose therapeutic strategy (non-drug + drug) | Management Plan: Choose therapeutic strategy (non-drug + drug) | Match | OK |
| 102 | Colombia | 2025-09-23 | Management Plan | Management Plan | Match | OK |
| 103 | México | 2025-09-30 | Management Plan | Management Plan | Match | OK |
| 104 | Perú | 2025-08-25 | Physiology and Function of the Atrioventricular Node | Evidence Synthesis / Research | Review | CHANGE: Non-canonical label ('Physiology and Function of the AV Node'); content is a synthesis of cardiac electrophysiology |
| 105 | México | 2026-01-06 | Work-up & Test Selection: Decide which tests/imaging to order | Evidence Synthesis / Research | Review | CHANGE: Classified as Work-up & Test Selection but requests recent articles on occupational health, not selection of tests to order |
| 106 | Perú | 2025-11-11 | Differential Diagnosis: Identify likely conditions explaining a presentation | Differential Diagnosis: Identify likely conditions explaining a presentation | Match | OK |
| 107 | Colombia | 2025-06-26 | Management Plan | Management Plan | Match | OK |
| 108 | México | 2025-08-19 | Evidence Synthesis / Research | Evidence Synthesis / Research | Match | OK |
| 109 | Colombia | 2025-11-06 | Prevention & Screening: Primary/secondary prevention | Prevention & Screening: Primary/secondary prevention | Match | OK |
| 110 | Argentina | 2025-11-20 | Differential Diagnosis: Identify likely conditions explaining a presentation | Differential Diagnosis: Identify likely conditions explaining a presentation | Match | OK |
| 111 | México | 2025-08-17 | Work-up & Test Selection | Work-up & Test Selection | Match | OK |
| 112 | México | 2026-01-05 | none_specified | Test Interpretation | Review | NOT CLASSIFIED IN DATABASE: interpretation of TSH >24 with elevated T4 values |
| 113 | México | 2025-12-08 | Patient Communication: Translate to lay language | Patient Communication: Translate to lay language | Match | OK |
| 114 | México | 2026-01-15 | Procedures & Techniques: How-to or indication for procedure | Procedures & Techniques: How-to or indication for procedure | Match | OK |
| 115 | Ecuador | 2025-07-19 | Management Plan | Management Plan | Match | OK |
| 116 | Perú | 2025-06-08 | Differential Diagnosis | Differential Diagnosis | Match | OK |
| 117 | Perú | 2025-08-20 | Management Plan | Management Plan | Match | OK |
| 118 | Colombia | 2025-12-09 | Differential Diagnosis: Identify likely conditions explaining a presentation | Differential Diagnosis: Identify likely conditions explaining a presentation | Match | OK |
| 119 | Colombia | 2026-01-05 | Procedures & Techniques: How-to or indication for procedure | Procedures & Techniques: How-to or indication for procedure | Match | OK |
| 120 | Colombia | 2025-08-12 | Pharmacotherapy | Pharmacotherapy | Match | OK |
| 121 | Perú | 2026-02-12 | Differential Diagnosis: Identify likely conditions explaining a presentation | Differential Diagnosis: Identify likely conditions explaining a presentation | Match | OK |
| 122 | México | 2026-05-22 | Evidence Synthesis / Research: Search for systematic reviews/RCTs | Evidence Synthesis / Research: Search for systematic reviews/RCTs | Match | OK |
| 123 | México | 2025-08-11 | Guideline Lookup | Guideline Lookup | Match | OK |
| 124 | Colombia | 2025-10-13 | Management Plan: Choose therapeutic strategy (non-drug + drug) | Management Plan: Choose therapeutic strategy (non-drug + drug) | Match | OK |
| 125 | México | 2026-01-28 | Work-up & Test Selection: Decide which tests/imaging to order | Work-up & Test Selection: Decide which tests/imaging to order | Match | OK |
| 126 | Perú | 2025-12-11 | Management Plan: Choose therapeutic strategy (non-drug + drug) | Management Plan: Choose therapeutic strategy (non-drug + drug) | Match | OK |
| 127 | México | 2025-07-23 | Differential Diagnosis | Differential Diagnosis | Match | OK |
| 128 | Chile | 2026-02-08 | Differential Diagnosis: Identify likely conditions explaining a presentation | Differential Diagnosis: Identify likely conditions explaining a presentation | Match | OK |
| 129 | Bolivia | 2025-07-04 | Differential Diagnosis | Differential Diagnosis | Match | OK |
| 130 | Bolivia | 2026-02-05 | Work-up & Test Selection: Decide which tests/imaging to order | Work-up & Test Selection: Decide which tests/imaging to order | Match | OK |
| 131 | Ecuador | 2025-08-15 | Management Plan: Choose therapeutic strategy (non-drug + drug) | Management Plan: Choose therapeutic strategy (non-drug + drug) | Match | OK |
| 132 | Perú | 2025-11-24 | Management Plan: Choose therapeutic strategy (non-drug + drug) | Management Plan: Choose therapeutic strategy (non-drug + drug) | Match | OK |
| 133 | Perú | 2026-02-10 | Work-up & Test Selection: Decide which tests/imaging to order | Pharmacotherapy | Review | CHANGE: Classified as Work-up & Test Selection but concerns indications, dosing, and safety of fluconazole in pediatrics |
| 134 | Perú | 2025-08-11 | Differential Diagnosis | Differential Diagnosis | Match | OK |
| 135 | México | 2025-11-18 | Work-up & Test Selection: Decide which tests/imaging to order | Work-up & Test Selection: Decide which tests/imaging to order | Match | OK |
| 136 | Nicaragua | 2025-09-08 | Management Plan | Management Plan | Match | OK |
| 137 | México | 2026-06-08 | Test Interpretation: Interpret numbers, visuals or cut-offs | Test Interpretation: Interpret numbers, visuals or cut-offs | Match | OK |
| 138 | México | 2025-11-25 | Management Plan: Choose therapeutic strategy (non-drug + drug) | Management Plan: Choose therapeutic strategy (non-drug + drug) | Match | OK |
| 139 | Perú | 2025-08-12 | Work-up & Test Selection; Guideline Lookup; Management Plan | Work-up & Test Selection; Guideline Lookup; Management Plan | Match | OK |
| 140 | Colombia | 2026-01-27 | none_specified | Summary / Overview | Review | NOT CLASSIFIED IN DATABASE: descriptive text on basic cardiac anatomy, educational context |
| 141 | México | 2025-08-12 | Differential Diagnosis | Differential Diagnosis | Match | OK |
| 142 | Colombia | 2025-09-15 | Test Interpretation | Test Interpretation | Match | OK |
| 143 | Ecuador | 2025-07-20 | Patient Communication | Patient Communication | Match | OK |
| 144 | México | 2025-09-29 | Management Plan | Management Plan | Match | OK |
| 145 | Perú | 2025-11-13 | Management Plan: Choose therapeutic strategy (non-drug + drug) | Management Plan: Choose therapeutic strategy (non-drug + drug) | Match | OK |
| 146 | Ecuador | 2025-09-17 | Test Interpretation | Test Interpretation | Match | OK |
| 147 | Colombia | 2025-10-16 | Differential Diagnosis: Identify likely conditions explaining a presentation | Evidence Synthesis / Research | Review | CHANGE: Classified as Differential Diagnosis but is a comparative anatomy table of shoulder and elbow, without a clinical presentation |
| 148 | Perú | 2025-11-13 | Evidence Synthesis / Research: Search for systematic reviews/RCTs | Evidence Synthesis / Research: Search for systematic reviews/RCTs | Match | OK |
| 149 | México | 2026-03-29 | Differential Diagnosis: Identify likely conditions explaining a presentation | Differential Diagnosis: Identify likely conditions explaining a presentation | Match | OK |
| 150 | Perú | 2025-11-20 | Evidence Synthesis / Research: Search for systematic reviews/RCTs | Evidence Synthesis / Research: Search for systematic reviews/RCTs | Match | OK |
| 151 | Colombia | 2025-08-21 | Pharmacotherapy | Pharmacotherapy | Match | OK |
| 152 | México | 2025-07-11 | Differential Diagnosis | Differential Diagnosis | Match | OK |
| 153 | México | 2025-12-08 | Management Plan: Choose therapeutic strategy (non-drug + drug) | Management Plan: Choose therapeutic strategy (non-drug + drug) | Match | OK |
| 154 | Venezuela | 2025-10-02 | Management Plan | Management Plan | Match | OK |
| 155 | Colombia | 2025-08-06 | Management Plan | Management Plan | Match | OK |
| 156 | Perú | 2026-06-01 | Differential Diagnosis: Identify likely conditions explaining a presentation | Differential Diagnosis: Identify likely conditions explaining a presentation | Match | OK |
| 157 | México | 2025-07-26 | Differential Diagnosis | Differential Diagnosis | Match | OK |
| 158 | México | 2026-02-05 | none_specified | Summary / Overview | Review | NOT CLASSIFIED IN DATABASE: academic description of glial cell types in the nervous system |
| 159 | Venezuela | 2025-09-23 | Differential Diagnosis | Differential Diagnosis | Match | OK |
| 160 | Ecuador | 2025-07-08 | Management Plan | Management Plan | Match | OK |
| 161 | Ecuador | 2025-08-16 | Test Interpretation | Test Interpretation | Match | OK |
| 162 | Colombia | 2025-09-18 | Test Interpretation | Test Interpretation | Match | OK |
| 163 | Perú | 2025-08-20 | Pharmacotherapy | Pharmacotherapy | Match | OK |
| 164 | México | 2025-09-01 | Pharmacotherapy | Pharmacotherapy | Match | OK |
| 165 | Perú | 2025-07-16 | Pharmacotherapy | Pharmacotherapy | Match | OK |
| 166 | Colombia | 2025-07-14 | Pharmacotherapy | Pharmacotherapy | Match | OK |
| 167 | Colombia | 2026-06-04 | Management Plan: Choose therapeutic strategy (non-drug + drug) | Patient Communication | Review | CHANGE: Classified as Management Plan but query is 'What does it mean' without clinical context; possible session start or ambiguous query |
| 168 | México | 2025-09-05 | Pharmacotherapy | Pharmacotherapy | Match | OK |
| 169 | Ecuador | 2025-11-24 | Management Plan: Choose therapeutic strategy (non-drug + drug) | Management Plan: Choose therapeutic strategy (non-drug + drug) | Match | OK |
| 170 | México | 2025-11-13 | Guideline Lookup: Locate authoritative guidance | Guideline Lookup: Locate authoritative guidance | Match | OK |
| 171 | Bolivia | 2026-02-02 | Differential Diagnosis: Identify likely conditions explaining a presentation | Differential Diagnosis: Identify likely conditions explaining a presentation | Match | OK |
| 172 | Ecuador | 2026-03-19 | Management Plan: Choose therapeutic strategy (non-drug + drug) | Management Plan: Choose therapeutic strategy (non-drug + drug) | Match | OK |
| 173 | Perú | 2025-08-27 | Management Plan | Management Plan | Match | OK |
| 174 | Perú | 2025-08-19 | Guideline Lookup | Guideline Lookup | Match | OK |
| 175 | México | 2025-09-26 | Work-up & Test Selection | Work-up & Test Selection | Match | OK |
| 176 | México | 2025-07-08 | nan | Summary / Overview | Review | NOT CLASSIFIED IN DATABASE: conceptual query on what echocardiography is |
| 177 | Bolivia | 2025-10-01 | Management Plan | Management Plan | Match | OK |
| 178 | México | 2025-06-19 | Management Plan | Management Plan | Match | OK |
| 179 | Bolivia | 2025-09-26 | Management Plan | Management Plan | Match | OK |
| 180 | Ecuador | 2025-12-03 | Work-up & Test Selection: Decide which tests/imaging to order | Pharmacotherapy | Review | CHANGE: Classified as Work-up & Test Selection but concerns indications and dosing of mometasone (nasal spray and topical) |
| 181 | Bolivia | 2025-11-19 | Work-up & Test Selection: Decide which tests/imaging to order | Guideline Lookup | Review | CHANGE: Classified as Work-up & Test Selection but concerns rejection criteria for samples submitted for bacteriological culture |
| 182 | México | 2025-11-22 | Test Interpretation: Interpret numbers, visuals or cut-offs | Test Interpretation: Interpret numbers, visuals or cut-offs | Match | OK |
| 183 | Ecuador | 2025-09-03 | Work-up & Test Selection | Work-up & Test Selection | Match | OK |
| 184 | Nicaragua | 2026-01-30 | Work-up & Test Selection: Decide which tests/imaging to order | Pharmacotherapy | Review | CHANGE: Classified as Work-up & Test Selection but compares albendazole vs. mebendazole for nematode treatment |
| 185 | Ecuador | 2025-06-08 | Test Interpretation | Test Interpretation | Match | OK |
| 186 | Bolivia | 2025-09-26 | Differential Diagnosis | Differential Diagnosis | Match | OK |
| 187 | Colombia | 2025-08-16 | Pharmacotherapy | Pharmacotherapy | Match | OK |
| 188 | Argentina | 2025-09-06 | General / Other | General / Other | Match | OK |
| 189 | Perú | 2026-02-05 | none_specified | Differential Diagnosis | Review | NOT CLASSIFIED IN DATABASE: infections as a precipitating factor for rejection in renal transplantation |
| 190 | Perú | 2025-08-23 | Pharmacotherapy | Pharmacotherapy | Match | OK |
| 191 | México | 2025-09-13 | Management Plan | Management Plan | Match | OK |
| 192 | México | 2025-08-20 | Test Interpretation | Test Interpretation | Match | OK |
| 193 | Venezuela | 2025-06-11 | Test Interpretation | Test Interpretation | Match | OK |
| 194 | Venezuela | 2025-06-18 | Pharmacotherapy | Pharmacotherapy | Match | OK |
| 195 | Chile | 2025-05-26 | Work-up & Test Selection | Work-up & Test Selection | Match | OK |
| 196 | Perú | 2025-12-06 | Work-up & Test Selection: Decide which tests/imaging to order | Pharmacotherapy | Review | CHANGE: Classified as Work-up & Test Selection but concerns meropenem dose adjustment in renal insufficiency |
| 197 | Perú | 2025-10-21 | Management Plan: Choose therapeutic strategy (non-drug + drug) | Management Plan: Choose therapeutic strategy (non-drug + drug) | Match | OK |
| 198 | Perú | 2025-09-03 | Evidence Synthesis / Research | Evidence Synthesis / Research | Match | OK |
| 199 | México | 2025-09-28 | Management Plan | Management Plan | Match | OK |
| 200 | Colombia | 2025-07-07 | Management Plan | Management Plan | Match | OK |

*NLP: natural language processing. Agreement: Match = NLP-assigned category confirmed by reviewer; Mismatch = category revised through team adjudication.*

**Supplementary Table S2. Monthly active physician counts and query volume by country (six high-volume countries).**

| **Month** | **México Q** | **MéxicoAP** | **Perú**  **Q** | **Perú AP** | **Colombia Q** | **Colombia AP** | **EcuadorQ** | **Ecuador AP** | **Bolivia**  **Q** | **Bolivia**  **AP** | **Venezuela Q** | **Venezuela AP** | **Total 6 Countries Q** |
| --- | --- | --- | --- | --- | --- | --- | --- | --- | --- | --- | --- | --- | --- |
| 2025-05 | 1882 | 148 | 1246 | 85 | 3912 | 284 | 990 | 81 | 1291 | 75 | 442 | 46 | 9763 |
| 2025-06 | 5662 | 561 | 3169 | 243 | 5643 | 513 | 2391 | 220 | 2666 | 191 | 1500 | 153 | 21031 |
| 2025-07 | 8275 | 711 | 5818 | 708 | 6376 | 648 | 3932 | 410 | 2829 | 328 | 1934 | 244 | 29164 |
| 2025-08 | 7018 | 725 | 5788 | 577 | 7385 | 734 | 2628 | 311 | 2018 | 254 | 1512 | 176 | 26349 |
| 2025-09 | 5680 | 532 | 7209 | 666 | 4867 | 480 | 2032 | 235 | 2264 | 213 | 1188 | 136 | 23240 |
| 2025-10 | 4487 | 319 | 4475 | 333 | 3353 | 272 | 1968 | 173 | 1566 | 152 | 990 | 98 | 16839 |
| 2025-11 | 4135 | 279 | 5427 | 310 | 3491 | 270 | 3102 | 156 | 2264 | 142 | 960 | 82 | 19379 |
| 2025-12 | 2913 | 215 | 4209 | 245 | 2395 | 180 | 1851 | 113 | 1481 | 106 | 735 | 68 | 13584 |
| 2026-01 | 3118 | 195 | 3653 | 205 | 2215 | 194 | 2202 | 123 | 1024 | 80 | 1033 | 86 | 13245 |
| 2026-02 | 3295 | 203 | 3106 | 182 | 2430 | 202 | 1848 | 118 | 1121 | 74 | 942 | 75 | 12742 |
| 2026-03 | 1380 | 162 | 1314 | 169 | 1115 | 153 | 756 | 86 | 536 | 82 | 368 | 62 | 5469 |
| 2026-04 | 1329 | 138 | 1559 | 165 | 1025 | 153 | 605 | 80 | 606 | 79 | 278 | 50 | 5402 |
| 2026-05 | 1238 | 124 | 1498 | 150 | 758 | 121 | 491 | 85 | 713 | 80 | 236 | 45 | 4934 |
| 2026-06 | 730 | 75 | 937 | 109 | 414 | 85 | 272 | 63 | 319 | 59 | 210 | 33 | 2882 |

*Q: queries; AP: Active physicians: physicians who generated at least one query in the calendar month. High-volume countries defined as those with >10,000 queries over the study period.*
